# Evaluating the biomedical and behavioural drivers of HIV-1 incidence decline in adolescent girls and young women in Uganda: A mathematical modelling study

**DOI:** 10.1101/2025.01.10.24319101

**Authors:** Adam Akullian, Victor Ssempijja, Daniel Breidenbecker, Fred Nalugoda, Gertrude Nakigozi, John Santelli, Philip Kreniske, Larry W Chang, Steven J. Reynolds, Robert Ssekubugu, Ronald H Gray, Maria J Wawer, Thomas C. Quinn, Ronald M. Galiwango, William J M Probert, Jeffrey W. Imai-Eaton, Oliver Ratmann, Christophe Fraser, Joseph Kagaayi, Godfrey Kigozi, M. Kate Grabowski, David Serwadda

**Affiliations:** Institute for Disease Modeling, Bill and Measslinda Gates Foundation; Clinical Monitoring Research Program Directorate, Frederick National Laboratory for Cancer Research; Rakai Health Sciences Program, Kalisizo, Uganda; Population and Family Health and Pediatrics, Columbia University; Graduate School of Public Health and Health Policy, City University of New York; Johns Hopkins School of Medicine and Bloomberg School of Public Health; Division of Intramural Research, National Institute of Allergy and Infectious Diseases, National Institutes of Health; Johns Hopkins University; Johns Hopkins Bloomberg School of Public Health; Department of Medicine, Johns Hopkins University School of Medicine; University of Oxford; Center for Communicable Disease Dynamics, Department of Epidemiology, Harvard T.H. Chan School of Public Health, Boston, MA, USA; MRC Centre for Global Infectious Disease Infectious Disease Analysis, School of Public Health, Imperial College London, London, United Kingdom; Department of Mathematics, Imperial College London; Li Ka Shing Centre for Health Information and Discovery, University of Oxford; Departments of Pathology and Epidemiology, Johns Hopkins University

## Abstract

**Background:** Recent declines in HIV incidence among adolescent girls and young women (AGYW) in Africa are often attributed to the expansion of biomedical interventions such as antiretroviral therapy and voluntary medical male circumcision. However, changes in sexual behaviour may also play a critical role. Understanding the relative contributions of these factors is essential for developing strategies to sustain and further reduce HIV transmission.

**Methods:** We conducted a mathematical modelling study of data from the Rakai Community Cohort Study (RCCS), an open, population-based cohort of 15- to 49-year-olds in 30 communities in Rakai, Uganda, to investigate the biomedical and behavioural drivers of HIV incidence decline in AGYW (15-24 years of age). We estimated changes in the HIV incidence rate between 2000-2019 using retrospective cohort data to validate our modelled incidence estimates. We ran modelled counterfactual scenarios to quantify the independent and combined effects (cumulative infections averted and difference in incidence rates) of antiretroviral therapy (ART), voluntary medical male circumcision (VMMC), and delays in age of first sex (AFS) over historical (between 2000-2020) and projected (between 2000-2050) time horizons.

**Findings:** Incidence in women 15-24 years of age declined by 83% between 2000-2019 (from 1.72 per 100 person-years in 2000 to 0.30 per 100 person-years in 2019), the largest reduction in incidence of all age groups of women. Increasing AFS over the last two decades (by 3 years in women and 2 years in men) was the largest contributor to incidence decline in women 15-19 years of age, averting 17% of cumulative infections between 2000-2020 and 37% between 2000-2050. Incidence in 15-19-year-old women was 69% lower in 2020 and 75% lower in 2050 compared to counterfactual scenarios without changes in AFS. ART scale-up contributed the most to incidence declines among women 20-24 years of age, averting 13% of infections between 2000-2020 and 43% of infections between 2000-2050. VMMC averted < 5% of infections in 15-24-year-olds to-date, with larger reductions in incidence between 2000-2050 in both 15-19 year-olds (13% reduction in cumulative infections) and 20-24 year-olds (22% of cumulative infections). ART, VMMC, and increasing AFS acted additively to reduce HIV incidence in AGYW, with little redundancy when combined.

**Interpretation:** Our results provide strong support for maintaining both the protective changes in sexual behaviours and effective biomedical interventions to sustain continued reductions in HIV incidence among AGYW.

## INTRODUCTION

Adolescent girls and young women (AGYW) in east and southern Africa have experienced among the highest rates of HIV acquisition in the world [1–3]. Several behavioural, biological, and socio-economic factors put young women at disproportionately high risk of HIV acquisition [4, 5]. These factors include higher per-sex-act risk of acquisition from increased biological susceptibility to HIV infection [6, 7], limited ability to negotiate safer sex and condom use [8], poverty and school dropout [3–5], gender-based violence [9], and higher prevalence of unsuppressed viral load in older male partners [10–12].

HIV incidence has declined among AGYW in the region over the last decade [13, 14], especially compared to that of older women, whose incidence has remained relatively stable in certain settings [14–17]. Multiple factors, including the scale-up of antiretroviral therapy (ART) [14, 18, 19] and voluntary medical male circumcision (VMMC) [19–21], as well as changes over time in behaviours that increase HIV risk [15], are likely contributors to incidence decline among AGYW [22], but no study has disentangled the independent effects of these hypothesised drivers.

Age at first sex (AFS) marks the initiation of exposure to STIs, including HIV and is associated with alarmingly high HIV risk in young African women [23]. Earlier initiation of sex is associated with higher risk of HIV acquisition [9, 24–28] via several socio-behavioural [9] and biological [29] pathways. Consequently, delaying sex protects against HIV infection by reducing the time spent sexually active, lowering the overall number of sexual partners [25], and reducing biological and behavioural susceptibility to infection. Several social and structural factors are hypothesized to drive delays in first sex, including rising socio-economic status (SES) and expanded educational opportunities for adolescents [30, 31]. It is unclear, however, to what extent recently observed delays in AFS in young adults [32] contribute to population-level declines in incidence among AGYW in the context of large-scale expansion of biomedical interventions like ART and VMMC.

Here we use a mathematical model calibrated to the Rakai Community Cohort Study, a population cohort study collecting prospective data on HIV incidences and determinants in southern Uganda since 1992, to quantify the independent and combined effects of treatment and prevention scale-up, and secular trends in AFS on historical and projected HIV incidence in AGYW. Our results have implications for continued investing in both socio-behavioural and biomedical interventions to reduce HIV incidence in AGYW.

## METHODS

### Study data

The Rakai Community Cohort Study is one of the longest-standing population-based HIV surveillance programs, following an open cohort of ∼20,000 adults from 30 continuously surveyed communities over a 30-year period. Details of the RCCS and survey methods are found elsewhere [15, 33]. Briefly, participants 15-49 years of age from a complete household census of 30 semi-urban and agrarian communities in four districts of southern Uganda were invited to participate in the longitudinal study. Those who provided written informed consent were interviewed every 1-2 years between 1999 and 2020, constituting 14 survey rounds (rounds 6-19). Self-reported data on demographics, socio-economic status, sexual risk behaviours, partnership attributes, ever ART use, and circumcision status (for men) were collected at each survey round. HIV testing was offered to all adult participants, and venous blood was obtained from those consenting to HIV testing. Laboratory methodology for HIV testing is described elsewhere [15]. The analysis was restricted to residents at the time of survey. The mean participation rate among adults eligible for the survey ranges from 59-67% across survey rounds, and uptake of HIV testing among those reached is high (∼99.7 tested upon offer of an HIV test across survey rounds) [17]. Similarly, self-reported ART coverage (percent of respondents who report ever having been on ART) was found to be similar in models adjusted for attributes associated with survey response [17], indicating low potential in selection bias.

### Estimation of HIV incidence, HIV prevalence, VMMC coverage, ART coverage, and age at first sex

HIV incidence was estimated from directly observed seroconversion among all participants with an HIV-negative test followed by at least one HIV test during a subsequent survey round. For those testing positive at a subsequent survey round, a seroconversion date was imputed randomly between the last negative and the first positive test, and a positive test was assigned to the survey round closest to that imputed date, and a negative status was assigned to the prior survey round. Follow-up was right censored at either the seroconversion date (for those testing positive) or last negative test (for those who remained serially negative). Those individuals with missing HIV status between two survey rounds with non-missing status had HIV status filled in according to the two non-missing tests. For example, an individual with positive status at rounds 6 and 10 and missing status in between had positive status filled in for rounds 7, 8, and 9, with a similar method applied to those with missing test data between two negative tests.

Person-years of follow-up and crude numbers of incident infections were calculated for each survey round for individuals included in the incidence cohort. Smoothed Poisson generalized additive models (GAMs) [34] with an offset for person-years at risk [35] and smooth terms for the main effects of age and survey round and an interaction between age and survey round were used to estimate sex-specific smoothed incidence rates and 95% confidence intervals. GAMs were fit with penalized, thin plate regression splines [34] (isotropic, locally weighted regression smoothers). The expected number of seroconversions *E(y_i_)* was estimated as the product of the Poisson-distributed seroconversion rate λ_i_ and person-time at risk *T_i_*. Linear combinations of 1- and 2-dimensional smooth terms, *f_1_,…,f_p_* were included as main effects in the model:

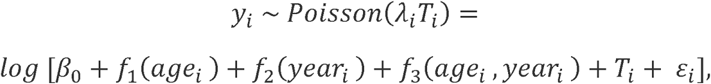

where β_0_ is an intercept, ε_i_ are identically distributed and independent (i.i.d.) errors, and *f_1_,…,f_p_* are smooth regression splines, defined as the sum of piece-wise polynomial basis functions *b_j_* and regression coefficients *Z_j_* with basis dimension *J*:

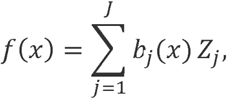

Generalized linear models (GLMs) were also fit with categorical terms for survey round and age group to estimate sex, age group, and round-specific incidence rates and 95% confidence bounds to compare with smoothed GAMs.

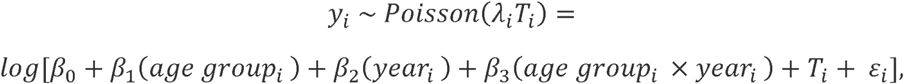

HIV prevalence was estimated cross-sectionally at each survey round using binomial GAMs with a logit link function:

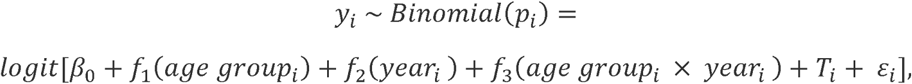

ART coverage (% on ART), male circumcision coverage (% circumcised), and prevalence of having initiated sex (% reporting ever having sex) were similarly estimated cross-sectionally at each survey round for all individuals with non-missing data on each indicator using the same binomial GAM specification as the HIV prevalence models.

### Mathematical modelling

Micro-simulation modelling was implemented using the epidemiological modelling software EMOD-HIV, an agent-based, stochastic, network transmission model described in detail at https://docs.idmod.org/projects/emod-hiv/en/latest and elsewhere [36]. EMOD simulates the natural history of HIV progression, considering the variable health states of individuals, such as CD4 cell count and WHO disease stage.

EMOD uses data on age-specific fertility, mortality, and sexual relationship formation [37], and models the cyclical flow of HIV-positive individuals through the HIV care cascade. ART and VMMC were allocated based on age, sex, and year-specific coverage estimates from cohort data. We estimated a per-contact ART efficacy of 92% [32], to reflect “real-world” barriers to suppression like adherence and undetected drug resistance. Other factors in our model that modify the transmission probability include condom usage, circumcision status of the male HIV-negative partner, age of the female HIV-negative partner (to capture increased biological susceptibility of younger women), and presence of an STI in either the HIV-negative or positive partner.

EMOD simulates the age, gender, and risk-group-specific sexual network using a sexual partner pair formation algorithm (PFA) described previously [37, 38]. The PFA provides a detailed representation of the age-specific structuring of heterosexual networks – a phenomenon that contributes to the age- and gender-specific patterns of HIV transmission [23, 37]. We capture the effects of delayed age at sexual debut on relationship formation by matching to survey data the percentage of individuals by age and sex available to form relationships on the network. In this way, changes in the proportion of individuals reaching sexual debut over time affect both the person-time exposed to HIV and the biological risk of acquisition among those delaying sex to older ages.

Models were calibrated to age-, gender-, and year-specific HIV prevalence and age-, gender-, and year-specific population numbers from the RCCS between 1999 and 2020. Cohort-based estimates of ART coverage, VMMC, and proportion initiating sex by age, sex, and year were specified as modelled inputs. PrEP was not included in the model because of limited availability of PrEP in the population and the uncertainty around its future scale-up in the population. A total of 100 best-fitting parameter sets were sampled from 6,600 simulations using roulette resampling in proportion to the likelihood. A summary of fixed parameter values and descriptions can be found in supplementary information (Tables S1 and S2).

### Model scenarios and outcomes

For historical (2000-2020) and future projections (2000-2050), we modelled HIV incidence per 100 person-years, cumulative infections averted, and cumulative % of infections averted, comparing a baseline scenario that captures the historical scale-up of all interventions and increasing in AFS to four counterfactual scenarios: 1) a scenario with no ART scale-up, 2) a scenario with no VMMC scale-up 3) a scenario with no changes in AFS and 4) a scenario with no interventions (including no changes in AFS). For each scenario, we held levels of each intervention and AFS constant from 2020-2050.

## Funding

This project has been funded in whole or in part with federal funds from the National Cancer Institute, National Institutes of Health, under Contract No. 75N91019D00024. The content of this publication does not necessarily reflect the views or policies of the Department of Health and Human Services, nor does mention of trade names, commercial products, or organizations imply endorsement by the U.S. Government. No conflicts of interest to declare. This work was supported by the National Institute of Allergy and Infectious Diseases (grant numbers U01AI100031, U01AI075115, R01AI110324, R01AI102939, R01AI128779, R01AI123002, and K01AI125086). This project was supported by the Division of Intramural Research, National Institute of Allergy and Infectious Diseases, National Institutes of Health (AI001040).

*This publication is based on research funded in whole or in part by the Gates Foundation, including models and data analysis performed by the Institute for Disease Modeling at the Bill & Melinda Gates Foundation*.

### Ethics

Ethical approval for the RCCS was obtained from the Uganda Virus Research Institute’s Research Ethics Committee (No: GC/127/19/11/137) and the Uganda National Council for Science and Technology (HS 540). Approval was also obtained from IRBs of collaborating institutions, including the Committee for Human Research at the Johns Hopkins University School of Public Health and School of Medicine (JHU NA_00069085) and the Western University Institutional Review Board. All participants provided written informed consent.

## RESULTS

### Study population

The population-based cohort of adult residents ages 15-49 included 40,801 participants (17,801 men and 23,000 women) surveyed over 14 rounds between 1999 and 2019. Mean age at first survey visit was 23.7 years in women and 24.1 years in men. Among women 15-24 included in the analysis of AGYW, at first survey visit, mean age was 18.8 years, 48.3% had ever been married, mean age at first sex was 15.7 years, and mean number of sex partners among those sexually active in the last year was 1.2.

### HIV incidence

The cohort used to estimate HIV incidence (those with a negative test followed by a subsequent test) included 22,080 participants aged 15-49, among whom 1,315 incident infections were observed (681 in women and 462 in men) in 75,442 person-years of follow-up in women and 65,003 person-years of follow-up in men. resulting in a crude incidence rate of 0.9/100py in women and 0.7/100py in men over the 20-year study period. A full enumeration of incident infections, person-years, incidence/100py, and 95% CI by age group and year for both men and women can be found in supplementary information table S1. Among women 15-24 years of age, 246 incident infections were observed in 22,615 person-years of observation, a crude incidence rate of 1.1/100py. HIV incidence per 100 person-years declined by 83% among women 15-24 years of age (from 1.72 per 100 py in round 7 to 0.30 per 100 py in round 19), 68% among women 25-34 years of age (from 1.15 per 100 py in round 7 to 0.37 per 100 py in round 19), and 63% among women 35-49 years of age (from 0.65 per 100 py in round 7 to 0.24 per 100 py in round 19) (Table 1). The larger relative decline in incidence in younger versus older women resulted in a shift to an older incidence age distribution in 2019 compared to 2000 (Figure 1). The age group with the highest incidence increased by 10 years over the 20-year study period, peaking among women 20-25 years in round 7 (year 2000) and among women 30-35 years of age in round 19 (year 2019).

**Fig 1.**
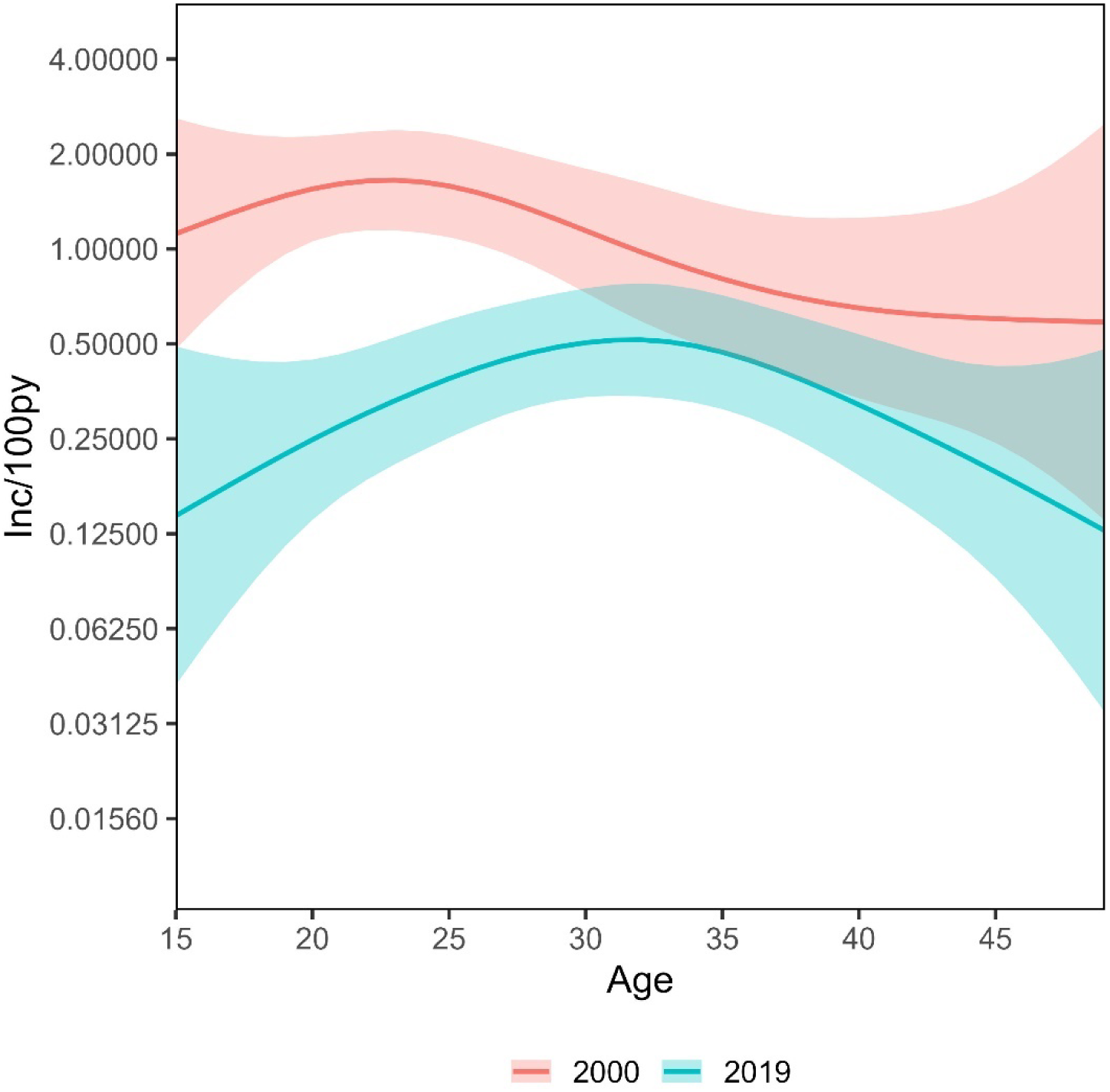
Smoothed age distribution of HIV incidence among women, estimated from empirical cohort data, between the first (year 2000) and last (year 2019) survey round. Note incidence is on a log2 scale.

**Table 1.** Modelled incidence rates, mean and 95% credible intervals, among AGYW by scenario, age group, and year.

| Scenario | Age | Year | Incidence per 100 py |  |  |
| --- | --- | --- | --- | --- | --- |
|  |  |  | mean | lb | ub |
| Baseline | 15-19 | 2020 | 0.26 | 0.15 | 0.40 |
|  |  | 2050 | 0.08 | 0.01 | 0.17 |
|  | 20-24 | 2020 | 0.73 | 0.44 | 1.01 |
|  |  | 2050 | 0.21 | 0.05 | 0.53 |
| No ART | 15-19 | 2020 | 0.57 | 0.38 | 0.79 |
|  |  | 2050 | 0.36 | 0.19 | 0.55 |
|  | 20-24 | 2020 | 1.66 | 1.13 | 2.29 |
|  |  | 2050 | 1.03 | 0.54 | 1.54 |
| No change in AFS | 15-19 | 2020 | 0.83 | 0.51 | 1.27 |
|  |  | 2050 | 0.31 | 0.08 | 0.77 |
|  | 20-24 | 2020 | 0.95 | 0.55 | 1.33 |
|  |  | 2050 | 0.38 | 0.11 | 0.73 |
| No VMMC | 15-19 | 2020 | 0.36 | 0.22 | 0.56 |
|  |  | 2050 | 0.17 | 0.05 | 0.33 |
|  | 20-24 | 2020 | 0.96 | 0.62 | 1.32 |
|  |  | 2050 | 0.52 | 0.16 | 0.92 |
| No Interventions | 15-19 | 2020 | 2.31 | 1.60 | 3.27 |
|  |  | 2050 | 2.70 | 1.65 | 3.98 |
|  | 20-24 | 2020 | 2.61 | 1.65 | 3.71 |
|  |  | 2050 | 3.09 | 1.63 | 4.63 |

### ART coverage, VMMC coverage, and percent ever initiating sex

Figure 2 shows temporal trends in ART coverage (% on ART), VMMC coverage (% circumcised), and the % of young men and women who reported ever initiating sex. ART began to scale-up after survey round 10 (year 2004), and steadily increased to survey round 19 (year 2019). ART scaled up earlier and to higher coverage in older men and women. By 2019, the highest coverage was in women 45-49 years of age (94%, 95% CI, 92-96%) and the lowest ART coverage in men 20-24 years of age (32%, 95% CI, 17-52%).

**Fig 2.**
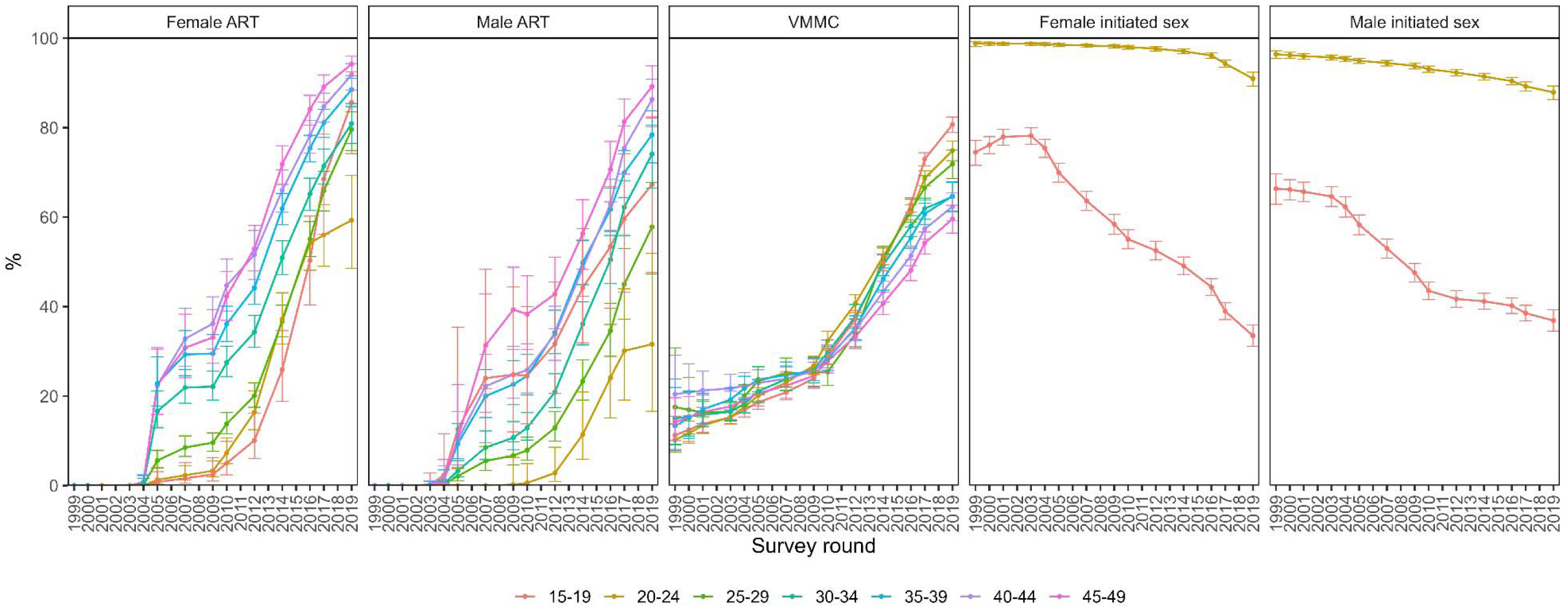
ART coverage (by age group and gender), VMMC coverage (by age group), and prevalence of initiated sex in those < 25 (by age group and gender) with 95% confidence bounds over time, estimated from smoothed generalized additive models with an interaction between year and age group.

The percentage of circumcised men increased steadily over the study period (Figure 2). VMMC scaled up slowly from 1999 to 2009, after which it climbed more steeply to round 2019. VMMC coverage was similar across all ages prior to 2010 and increased more rapidly in younger men thereafter. By 2019, 80% (95% CI, 79-82%) of men 15-19 were circumcised compared to 60% (95% CI, 56-63%) of men 45-49 years of age.

The proportion of men and women 15-24 years of age who reported ever having initiated sex declined over the course of the study (Figure 2)). Among women 15-19 years, half as many reported having initiated sex in 2019 compared to 1999 (from 80% to <40%). Similar declines were reported by young men. By 2019, similar proportions of young men and women reported having initiated sex.

### Mathematical modelling

Calibrated model fits to observed HIV prevalence by age, sex, and year are shown in supplementary information figures S2 and S3. Modelled HIV incidence per 100 person-years among women 15-24 between 2000-2020 had a similar trend as that of directly observed incidence estimated from the Rakai cohort, though modelled estimates tended to be higher than observed incidence (Figure 3). Modelled incidence declined from 2.1/100py (95% CI, 1.6-2.6/100py) in year 2000 to 0.47/100py (95% CI, 0.29-0.64/100py) in year 2020 and is projected to reach 0.2/100 py (95% CI, 0.07-0.39/100py) by 2040 with continuing declines to 2050.

**Fig 3.**
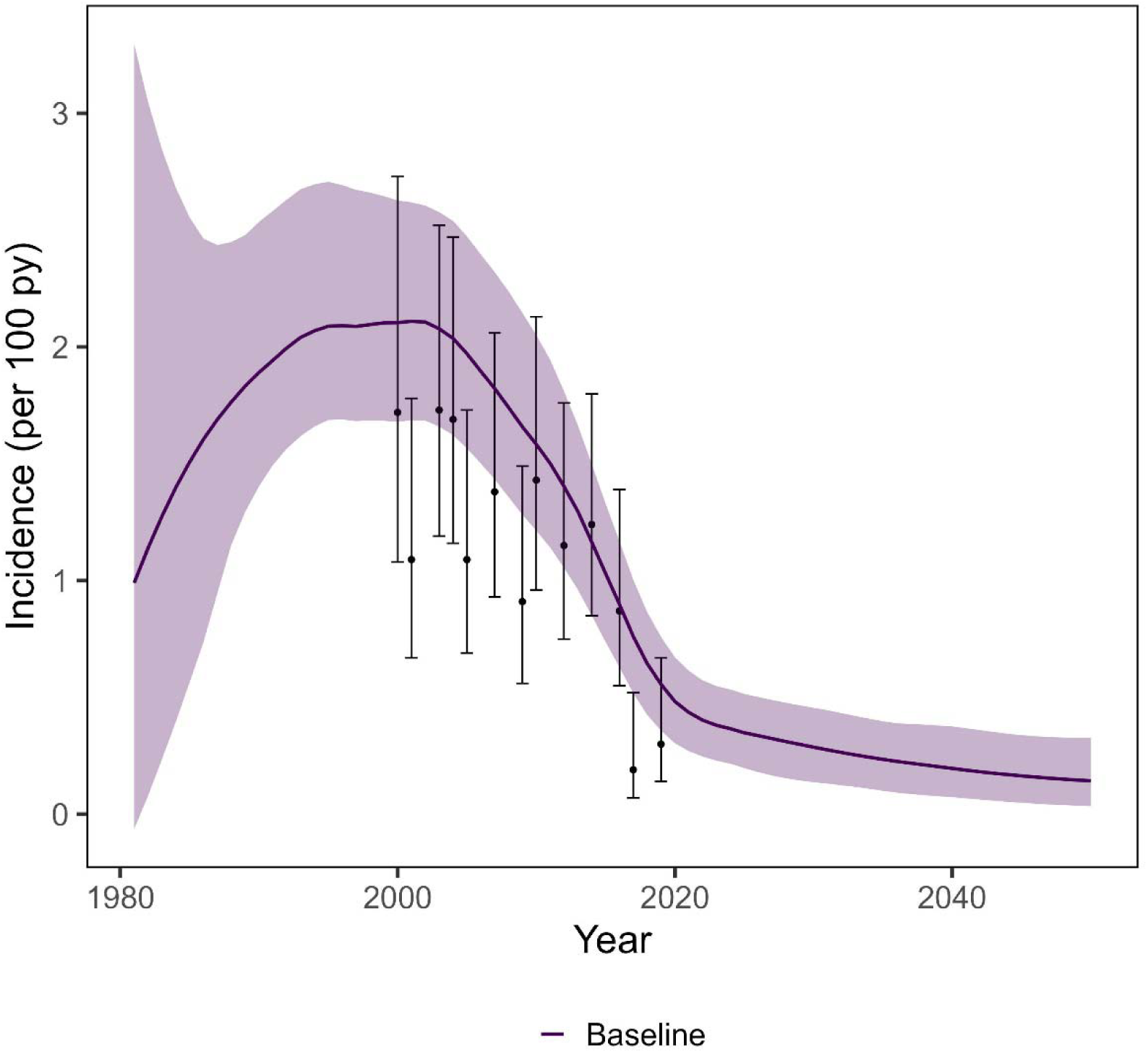
Comparison of modelled incidence (purple curve) and incidence observed in the Rakai cohort among women 15-24 years of age.

HIV incidence and cumulative infections averted are shown by age, gender, and year for each counterfactual scenario relative to the baseline scenario (Figure 4). Modelled increases in AFS (by 2 years in men and 3 years in women) (Figure 5a) had the largest effect on incidence in women 15-19 years of age, averting 17% of cumulative infections between 2000 and 2020 and 37% of cumulative infections between 2000 and 2050 (Figure 4). The incidence rate among 15-19 year olds in 2020 was 69% lower in the baseline (0.26/100py, 95% CI 0.15-0.40/100py) versus the counterfactual scenario with no change in AFS (0.83/100py, 95% CI 0.51-1.27/100 py) and in 2050 the incidence rate is projected to be 75% lower in the baseline (0.08/100py, 95% CI 0.01-0.17/100py) versus the counterfactual scenario with no change in AFS (0.31/100 py, 95% CI 0.08-0.77py) (Table 2). Among women 20-24 years of age, increases in AFS had no effect on cumulative incidence between 2000 and 2020, and averted 13% of cumulative infections by 2050. In women 20-24 years of age, incidence in 2050 is projected to be 45% lower in the baseline scenario (0.21/100py, 95% CI 0.05-0.53/100py) compared to the AFS counterfactual scenario (0.38/100py, 95% CI 0.11-0.73/100py) among women 20-24 years of age.

**Fig 4.**
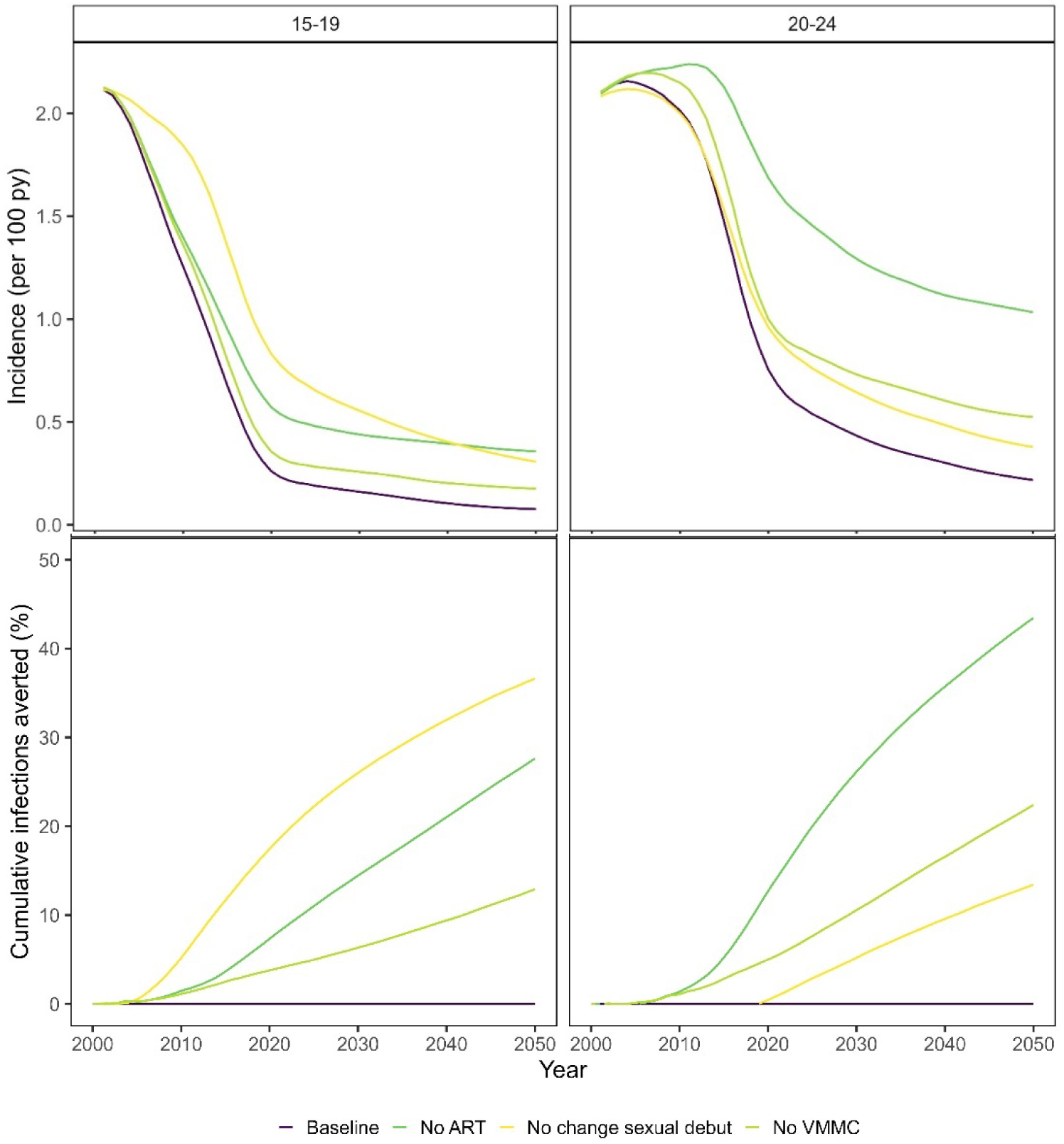
Modelled incidence rates (top panel) and cumulative infections averted (bottom panel) among women by scenario and age group. The percentage of cumulative infections averted compares cumulative infections in the counterfactual scenario (without the intervention) to the baseline scenario (with the intervention).

ART scale-up to date had a more moderate effect on incidence in women 15-19 to date and the largest effect of any intervention on incidence in women 20-24 years of age. ART scale-up averted 7% of infections in 15-19 y/o and 13% of infections in 20-24 y/o between 2000-2020. By 2050 ART scale-up is projected to avert 28% of infections in 15-19 y/o and 43% of infections in 20-24 y/o. In 2020 incidence in 15-19 y/o was 54% lower in the baseline scenario (0.26/100py, 95% CI 0.15-0.40) compared to the scenario without ART (0.57/100py, 95% CI 0.38-0.79), and by 2050 incidence in 15-19-year-olds is projected to be 78% lower in the baseline (0.08/100py, 95% CI 0.01-0.17) compared to incidence in the scenario without ART (0.36/100 py, 95% CI 0.19-0.55). VMMC scale-up to-date (Figure 2) had a small impact on incidence reduction in young women to date, averting 4% of cumulative infections in 15-19-year-olds and 5% of cumulative infections in women 20-24 years of age between 2000-2020, and a larger impact over longer time horizons: 13% of cumulative infections averted in 15-19-year-olds and 22% of cumulative infections averted in 20-24 y/o between 2000-2050.

Combined, all three scenarios (ART, VMMC, and increased AFS) reduced cumulative infections by 32% in 15-19 y/o and by 19% in 20-24 y/o between 2000-2020 and reduced cumulative infections by 74% in 15-19 y/o and 65% of cumulative infections in 20-24 y/o between 2000-2050. The combined effect of the three scenarios when modelled together was similar to the sum of the individual effects of each scenario for women 15-19 y/o over both time horizons and for 20-24 y/o between 2000-2020, indicating an additive effect between both interventions and increases in AFS. For women 20-24 y/o between 2000-2050, the combined effect for all scenarios was less than the sum of individual effects of each scenario, indicating some redundancy between ART, VMMC, and increasing AFS in that age group.

Modelled mean AFS increased by 2 years in men and 3 years in women between 2000 and 2020 (Figure 5a), shifting the age distribution of first sex older (Figure 5b). Increases in AFS changed the modelled structure of the sexual network. Under the baseline scenario, where AFS was delayed, men of all ages formed fewer total sexual relationships due to fewer sexual partnerships available with younger women (Figure 5d). However, in women, only younger women (< 23 years of age) formed fewer relationships (Figure 5d). Older women (23-31 years of age) formed *more* relationships, as men who were unable to form relationships with young women replaced those relationships with older women.

**Fig 5.**
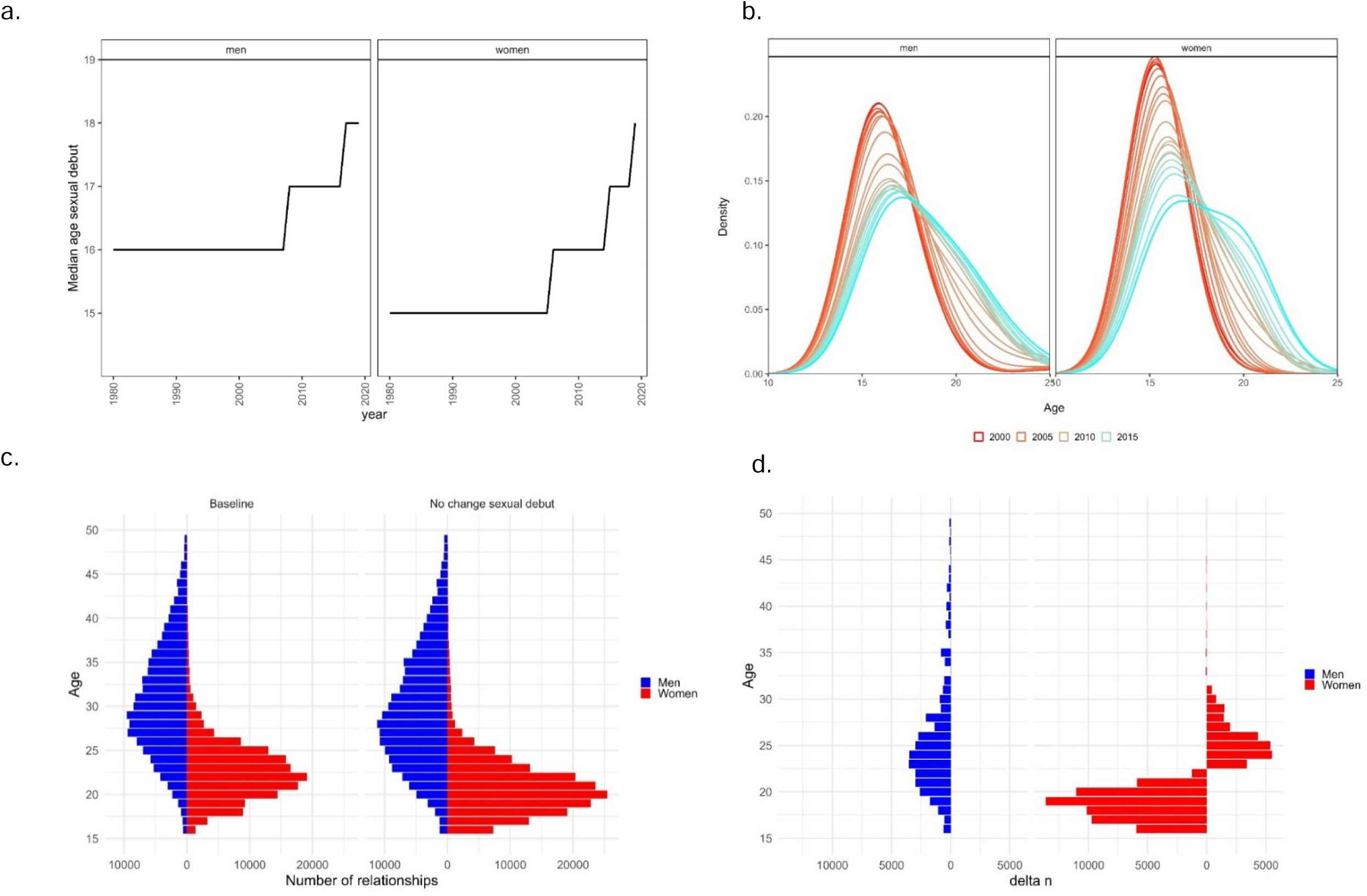
(a) Modelled median age at sexual debut by year, (b) distribution of age at sexual debut by year, (c) crude number of sexual relationships formed by age, gender, and scenario, and (d) difference in crude number of sexual relationships formed between baseline scenario and the scenario with no change in age at first sex (numbers to the left of zero indicate fewer relationships in the baseline scenario and numbers to the right of zero indicate more relationships in the baseline scenario).

## DISCUSSION

Through retrospective analysis of cohort data and mathematical modelling, we found that all of increasing AFS (by 2-3 years over the last decade), ART scale-up, and VMMC scale-up had substantial impacts on reducing HIV incidence among AGYW. Increases in AFS and ART scale-up were the largest contributors to incidence reductions in women 15-19 years of age, whereas ART alone contributed most to incidence decline in 20-24 years of age. The scale-up of VMMC impacted incidence over a longer time horizon, with a modest effect on historical incidence declines. These interventions, for the most part, acted additively, where the combined effects of all interventions were equivalent to the sum of their independent effects. Our results suggest that sustaining levels of these interventions will continue to reduce incidence in AGYW.

Considerable investments have been made in behavioural and biomedical prevention with the aim to reducing incidence in AGYW. The impact of these interventions on HIV risk in AGYW, however, has been mixed [39, 40]. The effectiveness of pre-exposure prophylaxis (PrEP) for HIV prevention in women under age 25 [41–43] has been suboptimal due to numerous barriers in product uptake and adherence. The efficacy of oral PrEP measured in clinical trials, for example, underperformed expectations due to low adherence in AGYW [44, 45]. Behaviour change interventions (BCIs) have also demonstrated mixed effectiveness in promoting safer sexual behaviours and reducing HIV risk [46]. Large-scale, multi-faceted social, economic, and biomedical interventions to reduce a constellation of contextual vulnerabilities and proximal risk factors to HIV have not shown impact [13, 47]. Sexual behaviour in young adults is complex and often negotiated in the context of imbalanced social, economic, and gender-based power dynamics within partnerships, thus challenging interventions that target individual-level behaviours.

Despite the mixed effectiveness of behavioural interventions targeted at AGYW at high risk of HIV acquisition, long-term trends in socio-economic status (SES) and educational attainment may reduce social and behavioural vulnerability to HIV over their life course. Educational attainment is proximally associated with several behavioural risk factors including older age at sexual debut, fewer lifetime number of partners, lower probability of having an older male partner, lower frequency of sex with a partner, and less frequent unprotected sex, which in turn lowers HIV risk [24, 28, 48, 49]. Rising SES and lower prevalence of orphanhood (resulting from a decline in HV mortality) in the Rakai Community Cohort study were associated with increases in school enrolment between 1994 and 2013, which in turn were associated with lower prevalence of HIV, lower adolescent pregnancy rates, and higher use of modern contraception [50]. The expansion of universal secondary education in Uganda in 2007 [51], which removed the barrier of school fees, may have also contributed to secular changes in sexual behaviour, and, in turn, to declining HIV risk in our study [31]. Whether this trend is observed in other settings can add to an evidence base supporting expanded access to secondary education for young women as part of HIV prevention programs.

We observed a larger effect of biomedical and behavioural interventions on incidence reduction in young women compared to older women. We found a disproportionately larger decline in incidence in women 15-24 years of age compared to older women, resulting in an age shift in incidence, a finding that has been confirmed from epidemiological [16] and phylogenetic data [17]. Reasons for age-heterogeneity in incidence decline are varied but include the larger relative reduction in HIV prevalence in younger versus older cohorts [16], targeted primary prevention to younger cohorts [52, 53], and disproportionately larger reductions in sexual risk behaviours, including delays in first sex, observed in younger cohorts [32]. Still, young adults in high-burden settings face challenges in achieving high coverage and effectiveness of treatment and prevention interventions that reduce risk in those populations. Most men living with HIV under the age of 25 and nearly 40% of young women living with HIV (20-24 y/o) reported no ART use, even under universal test and treat, a pattern consistently observed across populations in sub-Saharan Africa [54]. Given the importance of ART in reducing incidence in AGYW, targeted interventions towards increasing ART coverage in this age group, especially among men, could contribute to additional reductions in HIV transmission [17].

There are several epidemiological and model-based assumptions that may limit the interpretation of our analysis. First, we rely on self-report of ever having initiated sex, circumcision status, and ART use to parameterize our model and are subject to bias in level and trend over time. In modelling delays in first sex over time, we assume that older male partners who would otherwise form relationships with younger women (who are delaying sex) are replacing those relationships with older women. This can, in turn, increase the number of older individuals forming relationships on the network to offset the lower number of younger partners available for relationships. In this way, incidence may increase in older age groups, offsetting the incidence reduction from young women delaying sex. Previous modelling has shown that the effects of delaying sexual debut are sensitive to whether partnership replacement is assumed [55]. Further study is needed to understand how increasing AFS over time affects the structure of the sexual network, including any compensation for fewer young people available to form sexual relationships. These assumptions combined, however, would result in a lower impact of delayed AFS in our model, and our findings should thus be interpreted as a conservative estimate of those effects.

Second, incidence estimates for some years in our model were higher than cohort-based estimates. Our model may not have fully captured the risk distribution of the study population from which prevalence and incidence were estimated. One hypothesis is that higher-risk individuals may have been systematically missing from the population from which incidence was estimated. Incidence can only be directly observed in a resident population (who are present at multiple survey rounds). Mobile individuals, who tend to have higher HIV prevalence, would therefore be systematically excluded from the longitudinal cohort if they came into the cohort already positive. This would lead to a lower incidence in the population compared to our modelled estimates. Methods exist to correct for this bias by considering both left and right censoring using mechanistic modelling [56], though there are also limitations to this approach compared to directly observed methods. Strengths of our analysis include the ability to disentangle the hypothesised drivers of HIV incidence decline in AGYW, providing evidence that existing interventions act synergistically to impact both short and long-term incidence trajectories. We also dynamically model changes in sexual risk behaviour over time to match observed longitudinal trends, something that previous models have only done theoretically.

## Conclusion

Both protective changes in risk behaviour and scale-up of ART have contributed to incidence declines in AGYW (with VMMC acting over a longer time horizon), and if sustained, have the potential to continue to drive down infections in high-burden settings.

## Data Availability

All data produced in the present study are available upon reasonable request to the authors

## Supplementary information

**Fig S1.**
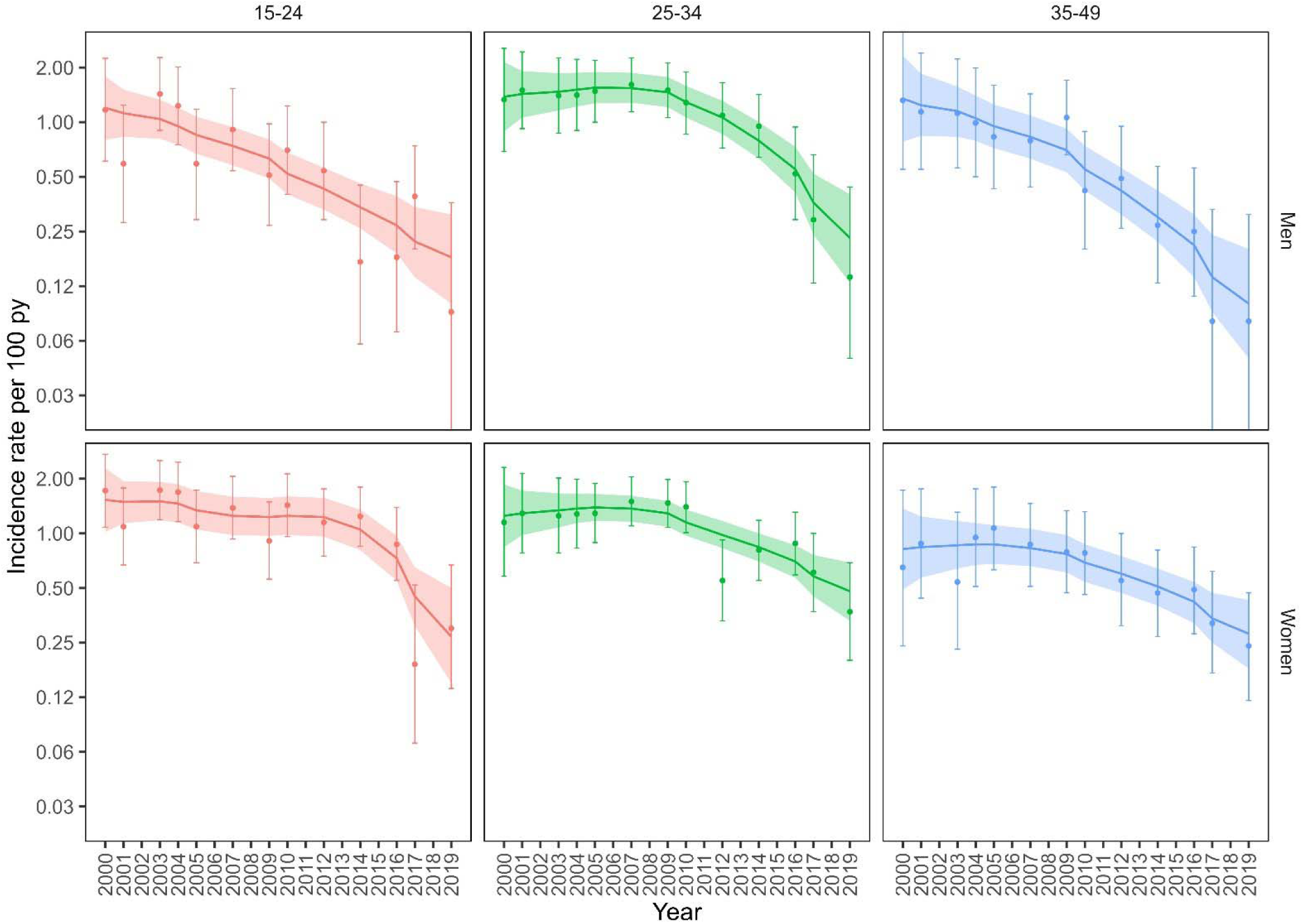
HIV incidence per 100 py by sex, year, and age group. Smoothed curves and 95% confidence intervals (ribbons) estimated from GAMs with smooth terms for year. Points and confidence intervals estimated from a GLM with categorical year. Note y-axis is on the log2 scale.

**Fig S2.**
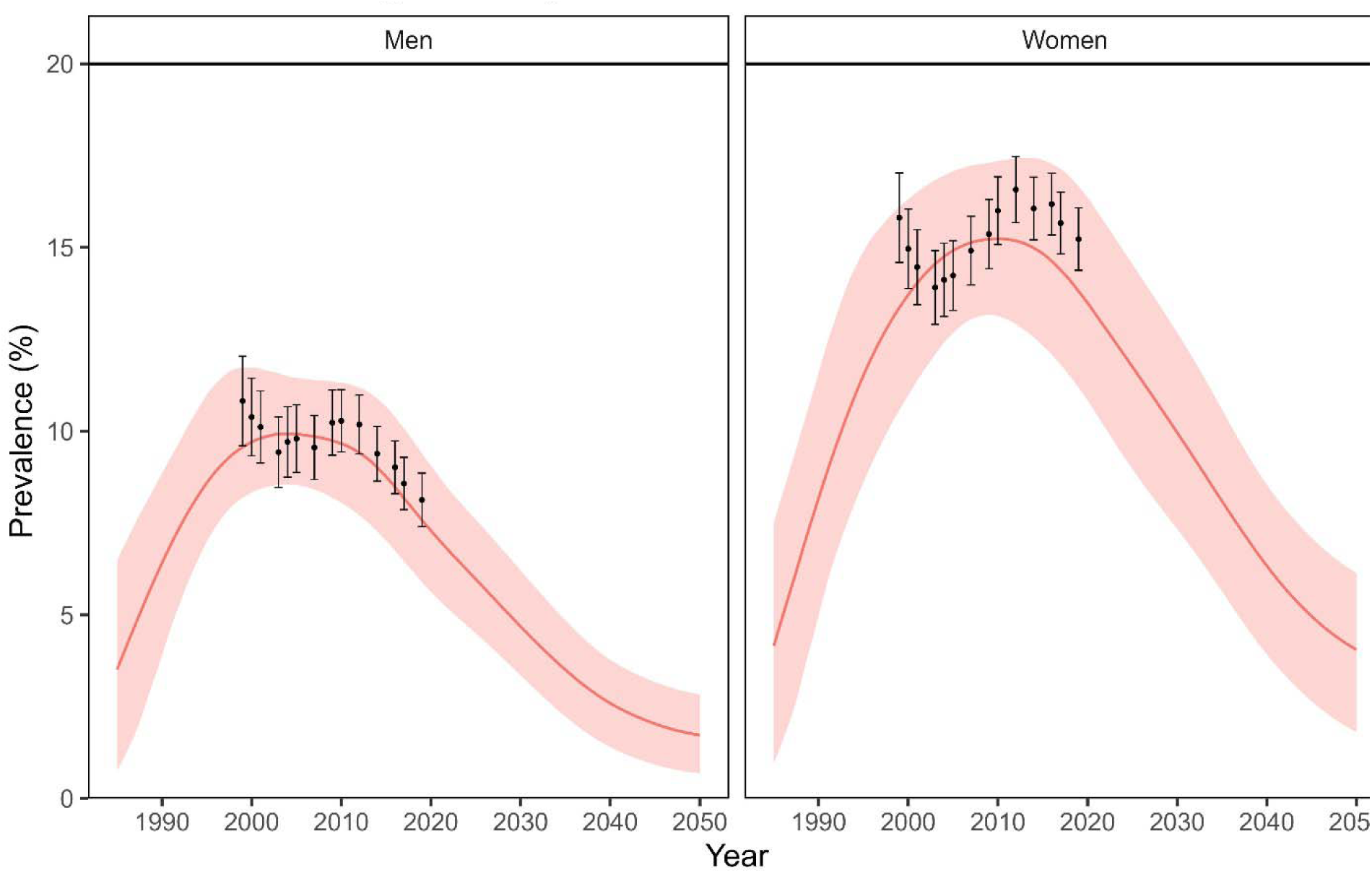
Modelled sex-, and year-specific prevalence and 95% credible interval (red curve) fit to observed prevalence in the Rakai cohort (black points with 95% confidence intervals) individuals 15-19 years of age.

**Fig S3.**
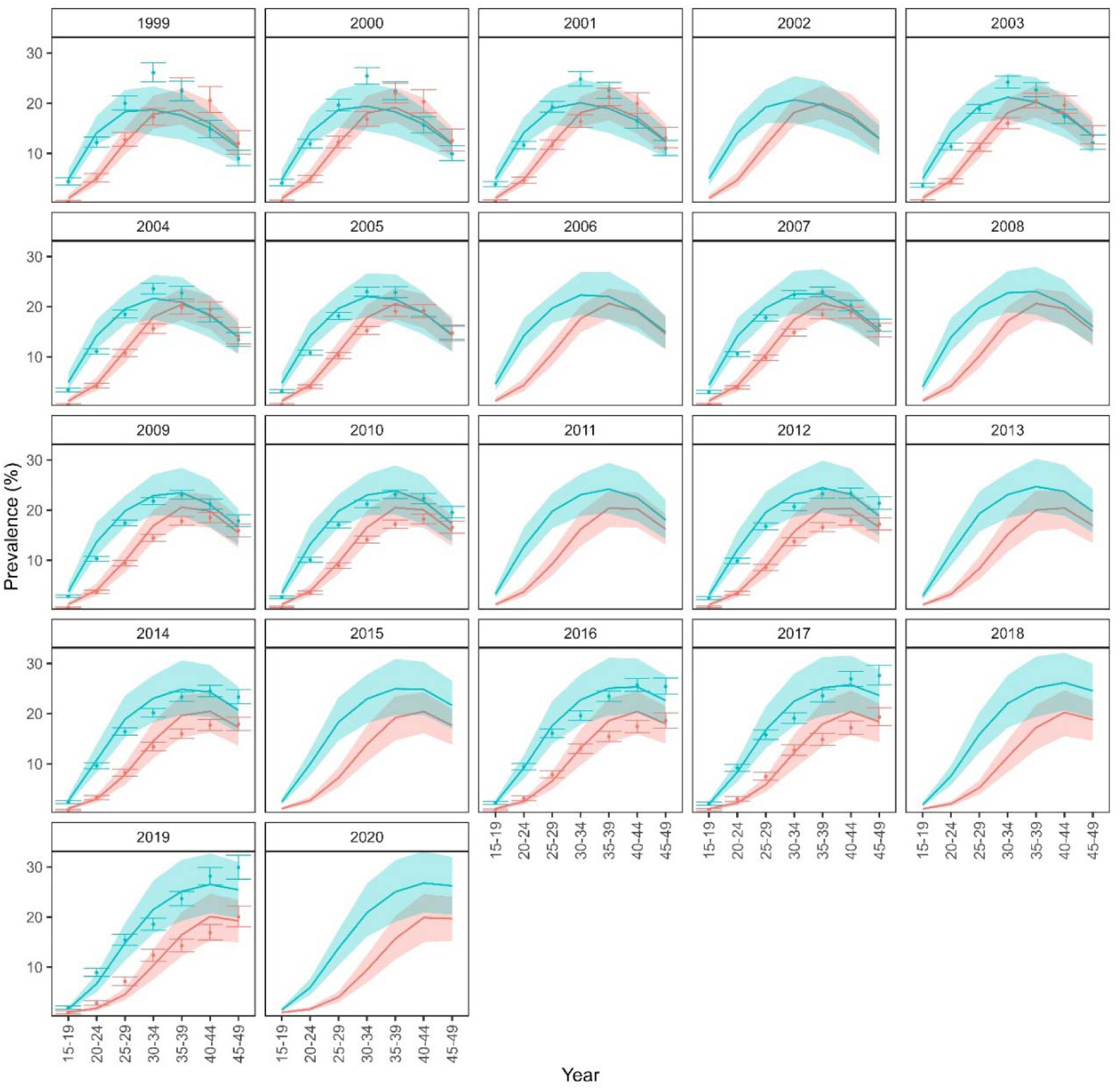
Modelled sex-, age-, and year-specific prevalence and 95% credible intervals (lines with shaded ribbons) fit to observed prevalence in the Rakai cohort (points and 95% confidence intervals.

---

**Table S1.**
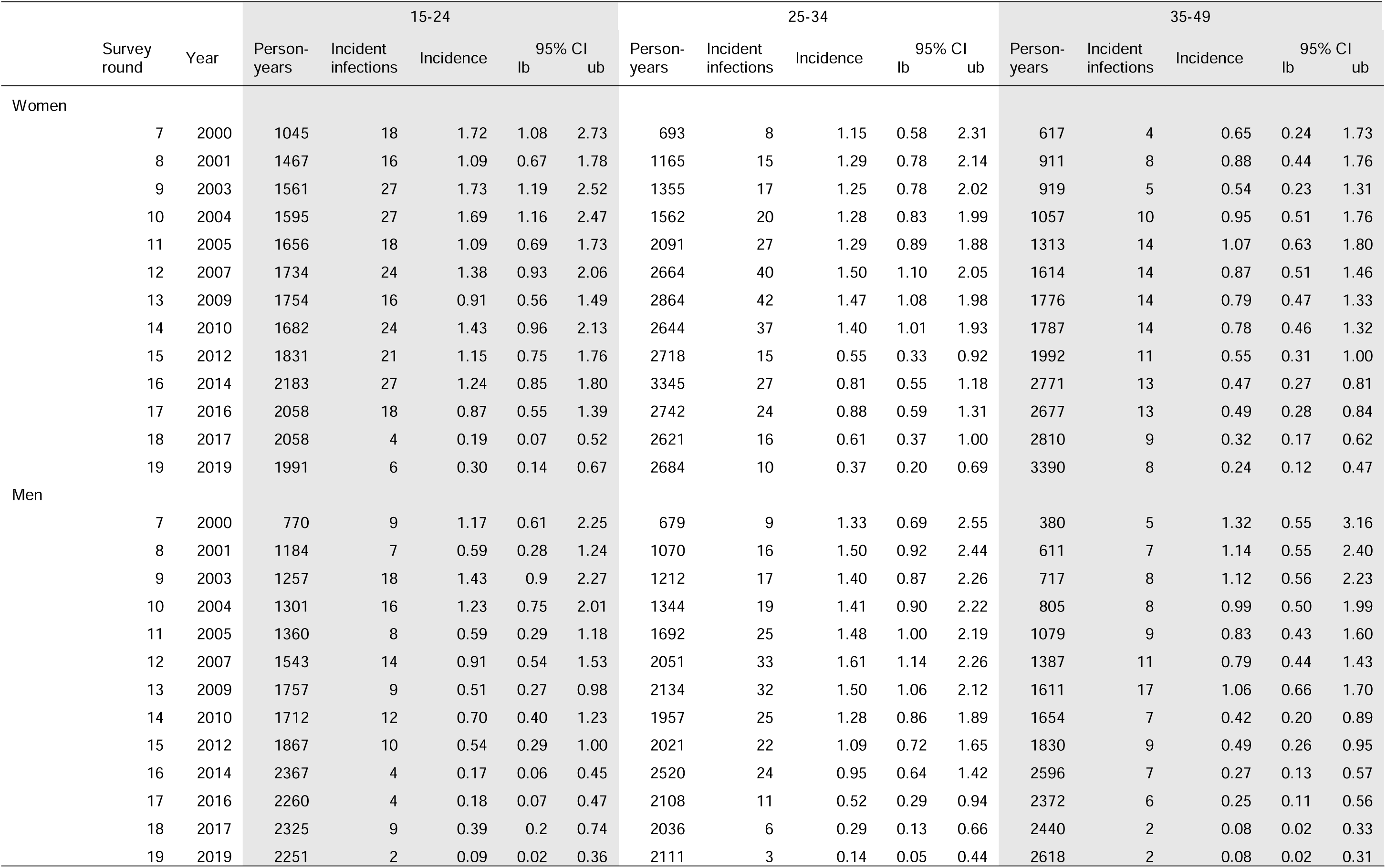
Person-years, incident infections, and incidence (per 100 py) estimates and 95% confidence intervals (CI) using GLMs by age group, sex, and survey round (with corresponding year).

**Table S2.** Median and interquartile range (IQR) of select calibrated parameters from 100 best-fitting parameter sets values.

|  | Parameter value quantile |  |  |
| --- | --- | --- | --- |
|  | 25% | 50% | 75% |
| Acute duration (months) | 3.99 | 4.24 | 4.70 |
| Acute stage multiplier on base infectivity | 10 | 10 | 18 |
| Base infectivity | 0.001142 | 0.001253 | 0.00141 |
| Circumcision efficacy | 0.62 | 0.66 | 0.69 |
| Initial proportion of low-risk individuals | 0.550 | 0.622 | 0.636 |
| Initial seeding proportion of infections into high-risk group (proportion) | 0.09 | 0.10 | 0.10 |
| Male to female infectivity multiplier for women 25 years and older | 1.62 | 1.81 | 2.21 |
| Male to female infectivity multiplier for women < 25 years | 3.58 | 4.59 | 6.36 |
| Mean drop out after ART initiation (days) | 1293 | 1324 | 1661 |
| Year HIV infections are seeded into high-risk group | 1980 | 1981 | 1983 |

**Table S3.** Select static model parameters used to fit the EMOD HIV transmission model to Rakai survey data (population, HIV prevalence, and ART coverage).

| Parameter | Description | Value / Median (IQR) |
| --- | --- | --- |
| AIDS_Duration_In_Months | The length of time, in months, prior to an AIDS-related death over which the AIDS_Stage_Infectivity_Multiplier is applied | 9 |
| AIDS_Stage_Infectivity_Multiplier | Multiplier acting on Base_Infectivity to determine the per-act transmission probability of an individual during AIDS stage | 4.5 |
| ART_CD4_at_Initiation_Saturating_Reduction_in_Mortality | The duration from ART enrollment to on-ART HIV-caused death increases with CD4 at ART initiation up to a threshold determined by this parameter value. | 350 |
| ART_Viral_Suppression_Multiplier | Multiplier acting on Base_Infectivity to determine the per-act transmission probability of a virally suppressed HIV+ individual. | 0.08 |
| CD4_At_Death_LogLogistic_Heterogeneity | The inverse shape parameter of a Weibull distribution that represents the at-death CD4 cell count. | 0.7 |
| CD4_At_Death_LogLogistic_Scale | The scale parameter of a Weibull distribution that represents the at-death CD4 cell count. | 2.96 |
| CD4_Post_Infection_Weibull_Heterogeneity | The inverse shape parameter of a Weibull distribution that represents the post-acute-infection CD4 cell count. | 0.2756 |
| CD4_Post_Infection_Weibull_Scale | The scale parameter of a Weibull distribution that represents the post-acute-infection CD4 cell count. | 560.43 |
| Coital_Act_Rate | Number of coital acts per day for all relationships except commercial ones | 0.33 |
| Coital_Act_Rate_Commercial | Number of coital acts per day for commercial relationships | 0.002739726 |
| Coital_Dilution_Factor_2_Partners | The multiplicative reduction in the coital act rate for all relationship types when an individual has exactly two current partners. Represents coital dilution. | 0.75 |
| Coital_Dilution_Factor_3_Partners | The multiplicative reduction in the coital act rate for all relationship types when an individual has exactly three current partners. Represents coital dilution. | 0.6 |
| Coital_Dilution_Factor_4_Plus_Partners | The multiplicative reduction in the coital act rate for all relationship types when an individual has exactly three current partners. Represents coital dilution. | 0.45 |
| Commercial_Condom_Max | The maximum asymptote for commercial relationships | 0.85 |
| Commercial_Condom_Mid | The year of the inflection point for commercial relationships | 1999.5 |
| Commercial_Condom_Min | The minimum asymptote of the probability of condom use per coital act for informal relationships for commercial relationships | 0.5 |
| Commercial_Condom_Rate | The rate proportional to the slope at the inflection point for commercial relationships | 1 |
| Commercial_Form_Rate | Exponentially distributed mean number new relationships formed per day for commercial relationships | 0.15 |
| Condom_Transmission_Blocking_Probability | The per-act multiplier of the transmission probability when a condom is used | 0.8 |
| Days_Between_Symptomatic_And_Death_Weibull_Heterogeneity | The time between the onset of AIDS symptoms and death is sampled from a Weibull distribution; this parameter governs the heterogeneity (inverse shape) of the Weibull. | 0.5 |
| Days_Between_Symptomatic_And_Death_Weibull_Scale | The time between the onset of AIDS symptoms and death is sampled from a Weibull distribution; this parameter governs the scale of the Weibull. | 618.341625 |
| HIV_Adult_Survival_Scale_Parameter_Intercept | Determines the intercept of the scale parameter for the Weibull distribution used to determine HIV survival time. Survival time with untreated HIV infection depends on the age of the individual at the time of infection, and is drawn from a Weibull distribution with shape parameter (see HIV_Adult_Survival_Shape_Parameter) and scale parameter. The scale parameter is allowed to vary linearly with age as follows<br>$\lambda = \text{HIV\_Adult\_Survival\_Scale\_Parameter\_Intercept} + \text{HIV\_Adult\_Survival\_Scale\_Parameter\_Slope} * \text{Age (in years)}$. | 21.182 |
| HIV_Adult_Survival_Scale_Parameter_Slope | This parameter determines the slope of the scale parameter for the Weibull distribution used to determine HIV survival time. | -0.2717 |
| HIV_Adult_Survival_Shape_Parameter | This parameter determines the shape of the Weibull distribution used to determine age-dependent survival time for individuals infected with HIV. | 2 |
| HIV_Age_Max_for_Adult_Age_Dependent_Survival | Survival time with untreated HIV infection depends on the age of the individual at the time of infection, and is drawn from a Weibull distribution with shape parameter and scale parameters (See HIV_Adult_Survival_Scale_Parameter_Intercept, HIV_Adult_Survival_Scale_Parameter_Slope, and HIV_Adult_Survival_Shape_Parameter). Although the scale parameter for survival time declines with age, it cannot become negative. To avoid negative survival times at older ages, this parameter, HIV_Age_Max_for_Adult_Age_Dependent_Survival, determines the age beyond which HIV survival is no longer affected by further aging. | 50 |
| HIV_Age_Max_for_Child_Survival_Function | The maximum age at which an individual's survival will be fit to the child survival function. If the value of this parameter falls between zero and the age of sexual debut, model results are not sensitive to this parameter as there is no mechanism for children to become infected between infancy and sexual debut. | 15 |
| HIV_Child_Survival_Rapid_Progressor_Fraction | The proportion of HIV-infected children who are rapid HIV progressors. | 0.57 |
| HIV_Child_Survival_Rapid_Progressor_Rate | The exponential decay rate, in years, describing the distribution of HIV survival for children who are rapid progressors. | 1.52 |
| HIV_Child_Survival_Slow_Progressor_Scale | The Weibull scale parameter describing the distribution of HIV survival for children who are slower progressors. | 16 |
| HIV_Child_Survival_Slow_Progressor_Shape | The Weibull shape parameter describing the distribution of HIV survival for children who are slower progressors. | 2.7 |
| Maternal_Infection_Transmission_Probability | The probability of transmission of infection from mother to infant at birth. | 0.3 |
| Maternal_Transmission_ART_Multiplier | The maternal transmission multiplier for on-ART mothers. | 0.03334 |
| Sexual_Debut_Age_Min | The minimum age at which individuals become eligible to form sexual relationships. | 13 |

